# Non-endoscopic screening for Barrett’s esophagus and Esophageal Adenocarcinoma in at risk Veterans

**DOI:** 10.1101/2024.03.15.24304354

**Authors:** Katarina B. Greer, Andrew E. Blum, Ashley L. Faulx, Erica M. Deming, Lauren L. Hricik, Hinnah Siddiqui, Brigid M. Wilson, Amitabh Chak

## Abstract

**Background:** While rates of Esophageal Adenocarcinoma (EAC) in the US continue to rise, many patients at risk of disease are not screened. EsoCheck (EC), a non-endoscopic esophageal balloon sampling device coupled with EsoGuard (EG), a DNA based screening assay, is an FDA-approved minimally invasive alternative to the traditional screening method of upper endoscopy.

**Aim:** Aim To prospectively determine the diagnostic accuracy, tolerance, and acceptability of the EC/EG test in a screening population.

**Methods:** We recruited Veterans who met the American College of Gastroenterology (ACG) Guideline criteria for endoscopic Barrett’s Esophagus (BE) and EAC screening at Louis Stokes Cleveland Veteran Affairs Medical Center. All study participants completed unsedated EC guided distal esophageal sampling followed by a sedated esophagogastroduodenoscopy (EGD). Diagnostic yield of the EG assay and EGD was recorded and used in calculation of sensitivity and specificity of EC/EG in prospective screening. The abbreviated Spielberger State-Trait Anxiety Inventory (STAI-6) questionnaire was administered before and after completion of EC. Overall tolerance of EC sampling was evaluated on a 10-point Likert scale.

**Results:** Results Esophageal cancer screening was accepted by 130/782 (16.6%) eligible veterans and we analyzed results of those who completed both screening tests (N = 124). Prevalence of BE/EAC among studied veterans was 12.9% (16/124), based on EGD. Sensitivity and specificity of EC/EG for EGD-detected BE/EAC were 92.9% (95% CI 66.1, 99.8) and 72.2% (95% CI 62.1, 80.8), respectively. Positive and negative predictive values were 32.5% (95% CI 18.6, 49.1) and 98.6% (95% CI 92.4, 100), respectively. Baseline STAI-6 scores were reflective of notable levels of anxiety among veterans in the peri-procedural setting. Mean post-procedure acceptability score for Esocheck test was 7.23 (SD 2.45).

**Conclusions:** Conclusions Our data suggest excellent sensitivity and negative predictive value of EC/EG in a screening population of veterans, making this modality a powerful screening tool for BE and EAC.

## Introduction

Esophageal adenocarcinoma (EAC) has increased more than 6-fold in the past four decades^1,2^. Twenty-one thousand five hundred sixty cases of EAC were diagnosed in the United States in 2023. The prognosis of patients with EAC is still poor with less than 22% of patients surviving beyond 5 years^2,3^.

BE remains the only known precursor of EAC and identification of BE is the only strategy for prevention or early detection of EAC. The major limitations of the current guidelines^4,5^ which recommend sedated EGD in patients with chronic gastroesophageal reflux disease (GERD) and additional BE risk factors, refractory gastroesophageal reflux disease (GERD), or alarm symptoms are clear. This strategy often fails to detect BE in patients whose symptoms are well controlled with either over the counter medications or physician prescribed therapies. It also fails to detect BE in subjects without diagnosis of GERD who comprise 40% of those that develop EAC^6,7^. Less than 5% of esophageal adenocarcinomas are caught diagnosed as early-stage lesions caught by surveillance of patients with previously detected BE^8^. Promising ablative non-surgical therapies that have been developed for early EAC and pre-malignant dysplastic BE^9^ over the past decade will have little impact on overall survival unless we develop more effective programs for identifying BE and early EAC in the general population. Effectiveness of early detection methods that are less costly, acceptable, accessible, and safer compared to screening EGD needs to be ascertained in clinical practice.

The veteran population is at increased risk for EAC and its precursor lesion, BE, due to increased prevalence of disease risk factors compared to the general population^10^. Prior study has shown that among Veterans diagnosed with esophageal adenocarcinoma, many had established risk factors for the disease but were not screened. Lack of BE screening in this veteran population represented the largest missed opportunity to reduce EAC mortality^11^.

We hypothesized that the incorporation of a non-endoscopic detection method in outpatient practice of a Veteran’s Affairs medical center could increase the positive predictive value of EGD and increase the detection of BE. The primary purpose of the study was to determine the accuracy of Esocheck/Esoguard (EC/EG) when used in prospective screening for BE and EAC among veterans. Secondary aims of this study were the following: 1) to determine the tolerability of the device when applied for screening and 2) to compare the diagnostic yield of 2 possible screening strategies: EGD only vs. Esocheck/Esoguard followed by EGD in patients who tested positive based on Esoguard assay.

## METHODS

### Patient population

Records of patients referred to gastroenterology for colon cancer screening and evaluation of upper gastrointestinal symptoms were reviewed to identify patients who met criteria for study enrollment. Eligible patients were adults between 40 and 85 years old who had history of symptomatic GERD and met criteria for upper endoscopic screening for BE based on current ACG and AGA guidelines (i.e. GERD symptoms plus three additional risk factors for BE including white race, obesity defined as body mass index (BMI) > 30 kg/m^2^, male gender, smoking history, family history of BE or EAC)^5,12^. Patients were excluded if they had known coagulopathy (INR > 1.5), esophageal varices, or significant dysphagia which would preclude them from swallowing the Esocheck device. All patients interested in screening completed baseline study questionnaires regarding demographics, risk factors, and history of reflux symptoms. Reflux symptom severity off medical therapy was assessed by GERD-Health Related Quality of Life (HRQL) questionnaire^13^.

### Procedure related anxiety

Anxiety related to study procedures was assessed based on Visual Analogue Scale (VAS) and State Trait Anxiety Inventory (STAI)-6 questionnaire^14^. On VAS, 0 represented “no anxiety” about the procedure, while 10 represented “most severe/worst anxiety”. STAI-6 is a shortened form of a validated 20 item questionnaire given to adults that assessed how strong a person’s feelings of anxiety are. It is designed to be administered in circumstances that prohibit the use of the full form such as pre-operative or ambulatory surgery settings. The questionnaire produces scores between 20 and 80 with higher scores indicating more severe anxiety.

### Study related procedures

The Esocheck was performed first, followed by screening upper endoscopy. Both procedures were performed on the same day. Sample collected from Esocheck was mailed to Lucid Diagnostics (Irvine, CA) for analysis and results were not available at the time of upper endoscopy. Endoscopists were blinded to Esocheck results and followed standard clinical practice regarding BE diagnosis. Routine biopsies of gastroesophageal junction were not performed; collection of tissue samples was reserved for patients in whom short or long segment esophagus was suspected based on white light and narrow band imaging inspection of the esophagus. If salmon colored mucosa was present, biopsies were obtained at 2-cm intervals along the entire length of the suspected Barrett’s esophagus following standard of care “Seattle” protocol.

### Sampling device

Esocheck is a 16X9mm encapsulated balloon device available commercially from Lucid Diagnostics (Lucid Technologies, New York, NY). The capsule is smaller than a multivitamin tablet and easy to swallow. The balloon hidden within the capsule is compliant to increase tolerability. It strategically inverts into the protective capsule when deflated, thus preventing sample contamination by cells within the proximal esophagus during device withdrawal. This feature allows to selectively sample the distal surface of the esophagus, without being overly abrasive.

### Molecular diagnostics

Samples were shipped to the Lucid Diagnostics CLIA-compliant lab for processing. DNA was extracted and stored at -70 degrees Celsius until analysis. EsoGuard assay utilizes bisulfite sequencing for detection of aberrant methylation in the vimentin and cyclin A1 genomic loci (mVim and mCCNA1, respectively). Samples are scored as VIM methylated if percent methylation is greater than 1.0%, and as CCNA1 methylated if percent methylation is greater than 0.5%. Samples were considered positive for the panel of mCCNA1 plus mVIM if either marker tested positive. When, for quality control purposes, a sample was run more than once, the results of the replicate runs were averaged. Molecular diagnostics lab was blinded to results of screening upper endoscopy.

### Timing of Esocheck and screening upper endoscopy

Esocheck was performed as the first study test. EGD was performed by gastroenterologists practicing in the Louis Stokes Cleveland VA Medical Center Endoscopy labs. The results of the molecular analysis were not available to the endoscopists or the study team at the time of screening upper endoscopy. Decision to collect tissue samples for diagnosis of Barrett’s esophagus was deferred to the endoscopist based on EGD findings.

### Statistical analysis

R statistical software (Vienna, Austria) was used to process the data. Categorical variables were summarized as the number of subjects and percentages. Continuous variables were summarized with means and standard deviations.

A two-by-two table was constructed comparing results from EC/EG screening and of EGD where EGD results were considered the true disease state. This table was used to determine sensitivity, specificity, positive predictive value, and negative predictive value of EC/EG and their exact 95% confidence intervals. This analysis was performed on the subset of subjects with both tests completed and with sufficient EC/EG sample to provide a result. In a sensitivity analysis, this analysis was repeated with insufficient sample EC/EG considered to be “positive” for the purposes of the screening as an EGD would be required in the absence of a negative result.

Logistic regressions were used to assess associations between number of risk factors and EGD result; and smoking status and EC/EG result. Pre- and post-procedure STAI-6 items and indices and visual anxiety scale were compared using paired t-tests. We assessed the correlation between pre-procedure STAI-6 and overall acceptability of the procedure.

Differences were considered significant for values of p<0.05.

Results obtained from screening procedures were used to compare two strategies: one where all patients go to screening EGD (strategy A) vs. a second strategy where patients have EC/EG, and then those who test positive proceed to diagnostic EGD (strategy B). Positive diagnostic yield of EGD and number of EGD procedures required to diagnose a given number of BE/EAC cases was determined for both strategies based on the prevalence observed in our analysis cohort and the positive predictive value (PPV) of EC/EG. A further sensitivity analysis altered strategy B such that patients with insufficient samples for EC/EG required a diagnostic EGD. The proportion of patients missed by the 2-step strategy was calculated from number of NP/n and its 95% confidence interval was determined with an exact binomial test.

## Study funding

Funding for this study was received by the Cleveland VA Research and Education Foundation on September 1st, 2021, from the Department of Defense. This trial has been registered with clinicaltrials.gov (NCT05210049).

## Results

Six thousand five hundred and eight patient charts with a diagnosis of chronic GERD have been **screened**, 782 were found eligible to fulfill study enrollment criteria and ***130 were enrolled*** into the study. Most common reasons for non-participation were completion of prior upper endoscopy outside of the VA system or unwillingness to participate in a research project.

Two patients signed consent and completed baseline questionnaires but were excluded from the study. The first enrolled patient was found to have esophageal varices, and the study team opted not to proceed with Esocheck. The diagnosis of cirrhosis with portal hypertension in this participant was an unexpected finding. The second was consented and filled out baseline study questionnaires during enrollment visit, but developed unrelated health issues between the time of consent and the appointment for study related procedures and did not complete Esocheck. Four patients (4/128, 3.1%) failed to swallow the Esocheck due to sensation of choking and fullness in the back of the throat. These patients still completed EGD and all study surveys and were screened for Barrett’s esophagus via upper endoscopy but were excluded from study analyses.

One hundred twenty-four patients completed both study tests. Of these, fourteen patients with Barrett’s esophagus and two patients with esophageal adenocarcinoma were identified for a prevalence of 12.9% (95% CI = 7.6%-20.1%). None of the patients with Barrett’s esophagus were found to have histologic dysplasia. Thirteen (13/124, 10.5%) swallowed the EC balloon, but an insufficient amount of DNA was collected to complete the Esoguard assay.

Baseline demographic characteristics of the enrolled patients are summarized in table 1. The mean age of the enrolled participants was 62.6 years (SD 10.7). Majority of the enrolled participants were white males. Six enrolled patients (4.8%) reported family history of Barrett’s esophagus or esophageal cancer. Majority of the enrolled participants were obese (70/124, 56.5%); mean body mass index (BMI) of study participants was 31.4kg/m^2^ (SD 6.2). Seventy eight percent of patients were using PPI at the time of enrollment into the study. The remainder had a diagnosis of chronic GERD In the medical record but used H2 blocker (i.e. Famotidine) or other over the counter medications for management of GERD symptoms. Mean GERD-HRQL score off PPI for enrolled patients was 21.2 (SD 11.7). Majority of enrolled participants admitted to active alcohol use. Seventy seven percent of patients reported current or prior history of smoking. Study participants reported that they smoked 0.77 (SD 1.04) packs per day.

**Table 1.** Baseline characteristics of patients enrolled into the study.

| Characteristic | Overall, N = 124<sup>I</sup> | GERD, N = 108<sup>I</sup> | BE or ECA, N = 16<sup>I</sup> |
| --- | --- | --- | --- |
| **Gender** |  |  |  |
| Male | 120 (97%) | 104 (96%) | 16 (100%) |
| Female | 4 (3.2%) | 4 (3.7%) | 0 (0%) |
| **Race** |  |  |  |
| White | 107 (86%) | 92 (85%) | 15 (94%) |
| Non-white | 17 (14%) | 16 (15%) | 1 (6.3%) |
| **Age** | 63 (11) | 62 (11) | 65 (9) |
| **BMI** | 31.4 (6.2) | 31.6 (6.2) | 30.2 (5.7) |
| **Smoking** |  |  |  |
| Never | 27 (22%) | 25 (23%) | 2 (13%) |
| Current | 41 (33%) | 35 (33%) | 6 (38%) |
| Past | 55 (45%) | 47 (44%) | 8 (50%) |
| Unknown | 1 | 1 | 0 |
| **Packs per day smoked** | 0.77 (1.04) | 0.77 (1.09) | 0.74 (0.64) |
| Unknown | 2 | 1 | 1 |
| **Family history of BE/EAC** |  |  |  |
| Yes | 6 (4.8%) | 5 (4.6%) | 1 (6.3%) |
| No | 118 (95%) | 103 (95%) | 15 (94%) |
| **Alcohol use** |  |  |  |
| Yes | 67 (54%) | 58 (54%) | 9 (56%) |
| No | 56 (46%) | 49 (46%) | 7 (44%) |
| Unknown | 1 | 1 | 0 |
| **Alcohol Servings/Week** | 1.05 (1.22) | 1.03 (1.19) | 1.19 (1.42) |
| Unknown | 1 | 1 | 0 |
| **Number of risk factors** | 4.11 (0.82) | 4.07 (0.81) | 4.38 (0.89) |
| **GERD HRQL score** | 21 (12) | 21 (11) | 22 (15) |
| **Pre-procedure STAI-6 score** | 36 (12) | 37 (12) | 30 (12) |
| Unknown | 1 | 1 | 0 |
| **Post-procedure acceptability score** | 7.23 (2.45) | 7.31 (2.39) | 6.63 (2.85) |

<sup>I</sup> n (%); Mean (SD)

The mean number of risk factors among enrolled patients was 4.1 (SD 0.8). One of the enrolled participants diagnosed with BE/EAC had a family history of the condition. Among subjects who were newly diagnosed with BE/EAC, there was following distribution of number of risk factors for the condition: 2 patients had 3 risk factors, 8 patients had 4 risk factors, 4 patients had 5 risk factors, and 2 patients had 6 risk factors. Each additional risk factor showed nonsignificant increase in likelihood of BE diagnosis (OR=1.57, 95% CI = (0.83, 3.10), p = 0.17).

Most of the patients diagnosed with Barrett’s esophagus had short segment disease. Six patients had long segment BE (Table 2). 8/14 (57.1%) patients diagnosed with BE had not been referred for screening by their providers prior to enrollment despite having multiple risk factors for the condition.

**Table 2.**
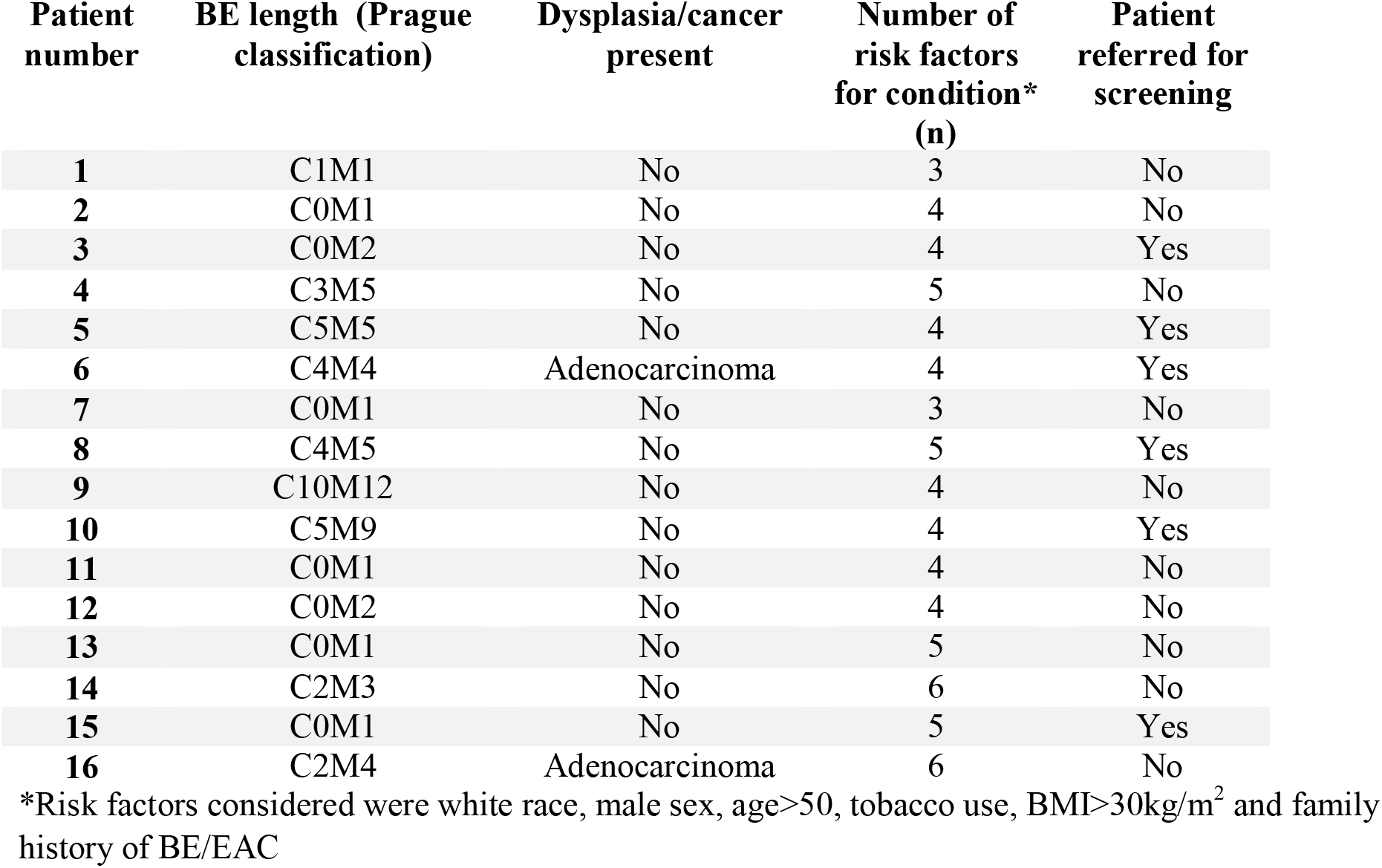
Characteristics of patients diagnosed with Barrett’s esophagus or Esophageal Adenocarcinoma.

Endoscopic findings in patients with false positive EC/EG individuals included normal mucosa (21/27, 77.8%), reflux esophagitis (5/27, 18.5%) or Candida esophagitis (1/27, 3.7%). Smoking status (never vs. current vs. former) was not predictive of EG result (likelihood ratio test p=0.49). Current smoking was reported in 14/40 (35%) of positive EG assays and 8/27 (29.6%) of false positive EG test results.

In our prospectively recruited study of Veterans with chronic GERD, the prevalence of BE was 12.9% (95%CI 7.6, 20.1). When patients with both complete diagnostic tests and sufficient DNA samples for EC/EG were included in the analysis (n= 111), EC/EG showed a sensitivity of 92.9% (95% CI 66.1, 99.8), specificity of 72.2% (95% CI 62.1, 80.8), positive predictive value of 32.5% (95% CI 18.6, 49.1) of and a negative predictive value of 98.6% (95% CI 92.4, 100). Diagnostic accuracy of EC/EG was 74.8% (95% CI 65.6, 82.5). When participants who had insufficient DNA samples were included in the analysis (n=124) as presumed EC/EG positive for purposes of the screening process, the sensitivity of EC/EG was 93.8% (95%CI 69.8, 99.8), and negative predictive value of the test was 98.6%, 95% CI 92.4, 100).

Baseline STAI-6 scores were reflective of notable levels of anxiety among veterans in the peri-procedural setting. Mean pre-procedure STAI-6 score for veterans was 35.7 (SD 12.5). Mean difference within subject between pre and post procedure STAI-6 scores was a decrease of 1.5 (95% CI -0.8, 3.8, p=0.20). In the individual domains that create the STAI-6 score, pre-test and post-test results yielded statistically significant drop in the category of worry (Figure 2). Mean pre-procedure “worry” score was 1.76 (SD 0.95) while the post-procedure “worry” score was 1.40 (SD 0.81). In the remaining categories of feeling calm, tense, upset, relaxed, and content there were no statistically significant changes in pre- and post-procedure test values. Pre-procedure STAI-6 scores did not significantly correlate with overall satisfaction with the procedure although higher baseline anxiety showed statistically insignificant negative correlation with overall procedure satisfaction (Pearson rho =-0.15, 95% CI -0.31, 0.03, p=0.11).

**Figure 1.**
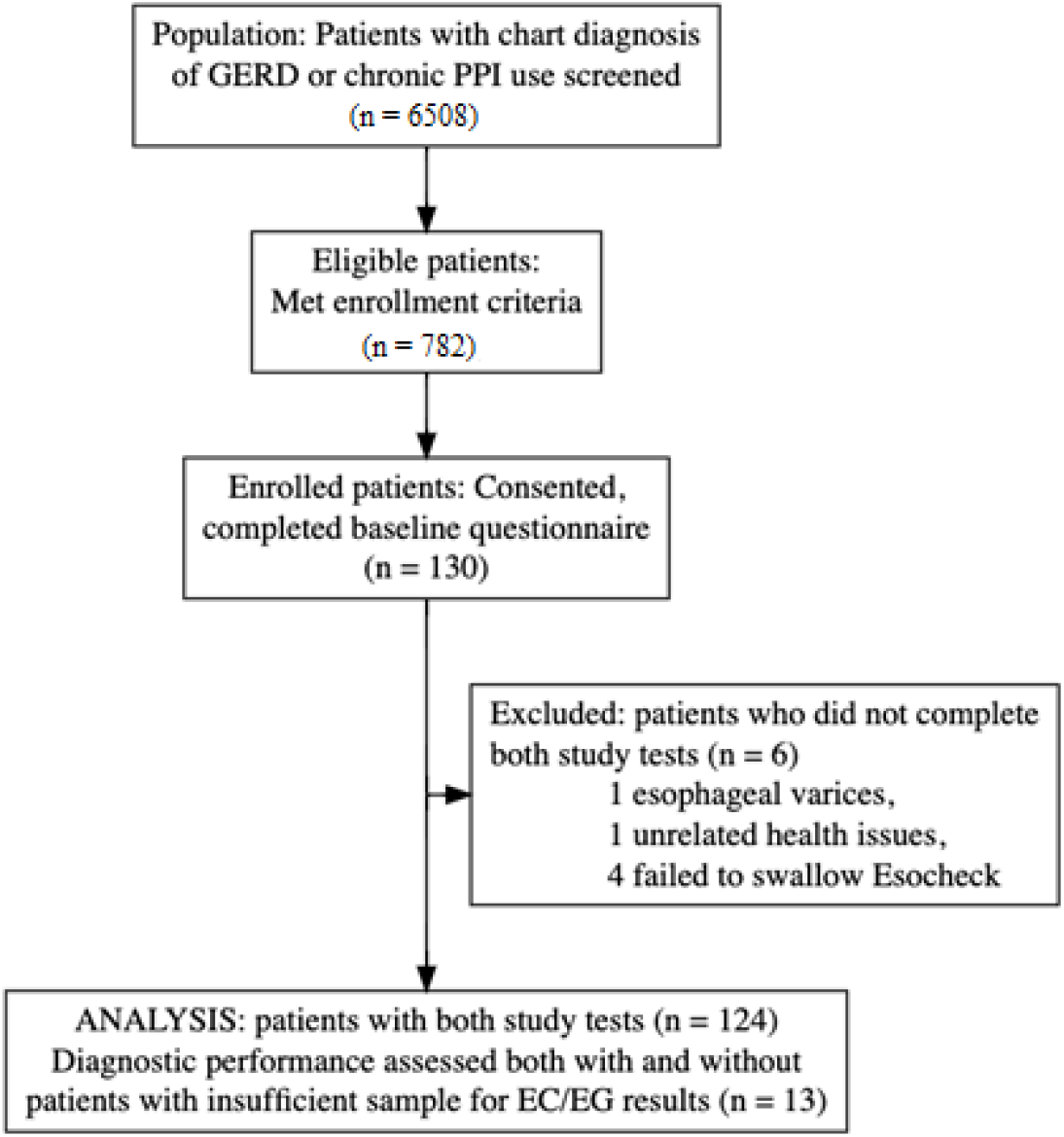
Flow chart of patient recruitment into the study (numbers need updated)

**Figure 2.**
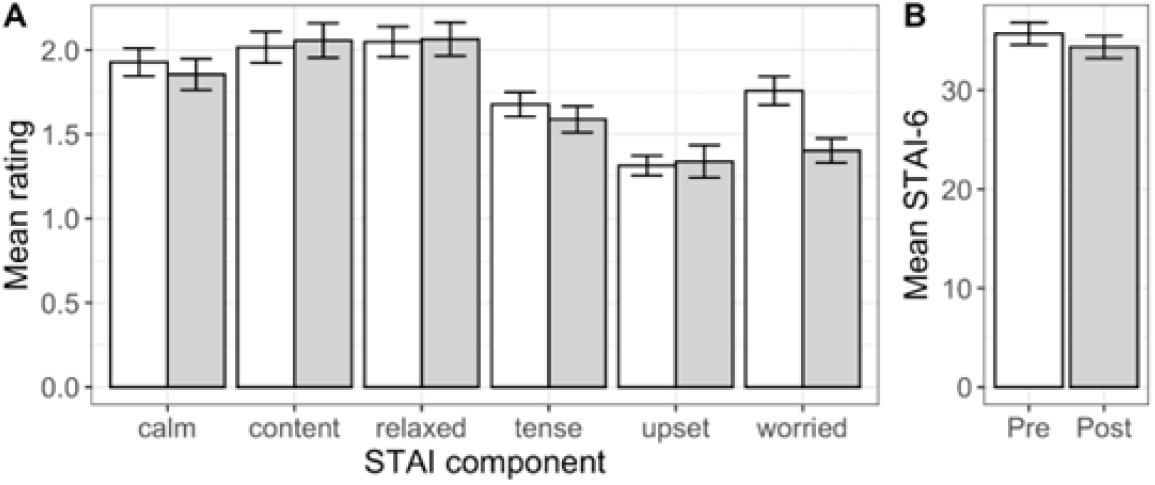
Procedure related anxiety scores before and after Esocheck test. Panel A shows individual STAI-6 domain scores. Panel B shows STAI-6 composite scores. Mean item or index presented with standard errors; pre-procedure shown as white bars; post-procedure shown as gray bars.

**Figure 3.**
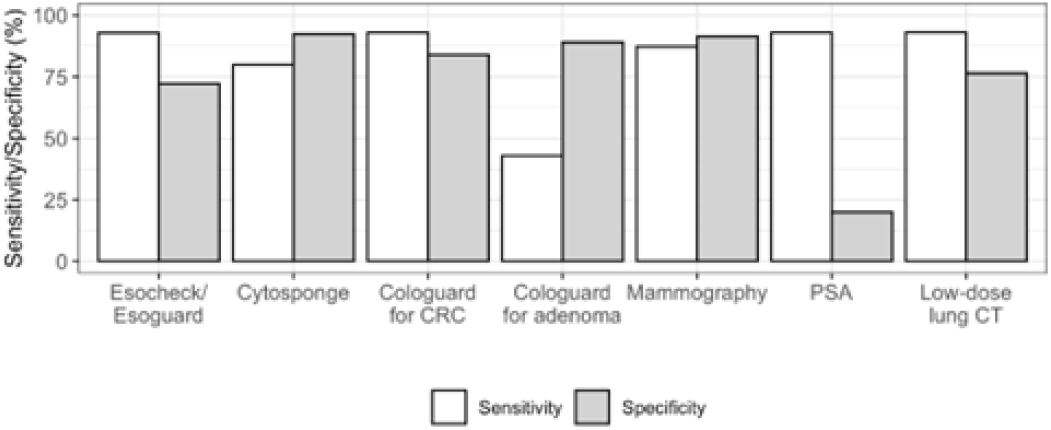
Comparison of Esocheck/Esoguard sensitivity and specificity to other screening tests for esophageal, colorectal, breast, prostate and lung cancer.

Based on visual anxiety scale (VAS) of 0-10 where 0 and 10 represented no anxiety and worst possible anxiety respectively, mean pre-procedure VAS score was 2.4 (SD 2.4) and post-procedure VAS score was 1.5 (SD 2.2). Mean difference in pre- and post-procedure VAS scores was statistically significant (0.8, 95% CI 0.4, 1.3, p<0.001), demonstrating decrease in anxiety after completion of EC.

Overall procedure acceptability was rated on a visual scale of 0-10 with 0 being the worst experience ever and 10 being the best experience. Mean post-procedure acceptability score for EC test was 7.23 (SD 2.45).

Assuming the prevalence observed in our studied cohort when comparing the use of EGD alone (Strategy A) with hypothetical strategy of ordering EC first and following up on all positive EG assays by EGD (Strategy B), strategy B decreased the number of EGDs necessary to diagnose the same number of BE/EAC by 60%. If we also considered a real-life scenario where EGD would be completed for all positive or insufficient sample Esoguard assays, the number of EGDs performed would be reduced by 53%. The number of cases that EC/EG missed which tested positive for BE/EAC by EGD was calculated to be 9.0 (95% CI = 0.2, 49) per 1,000.

## Discussion

This study describes the diagnostic accuracy of the EsoCheck device, a non-endoscopic screening method for BE and EAC, in real life clinical settings and prospectively recruited patients. With its strong negative predictive power, this screening modality could be a first line tool available to a greater number of patients. Data from this test supports the notion that Esocheck could be performed as a triaging test to increase the yield of diagnostic upper endoscopy 2.5-fold. The sensitivity of Esocheck at 92% compares well with non-invasive tests for cancers where USPTF recommends screening and which are already in use^15,16,17^. In this small study two cases of EAC were detected and none of the cases diagnosed with BE harbored dysplasia. The study was not powered to prospectively determine Esocheck diagnostic accuracy for subgroups of non-dysplastic, dysplastic BE and EAC. This data is reported for this device in development studies but not available for our study population.

When compared to the Cytosponge-TFF3, another non-endoscopic screening device for EAC and BE, the Cytosponge exhibited a lower range of sensitivity of 79.5-87.2% depending on the length of the lesion but it had a higher specificity of 92.4%^18^. This discrepancy in sensitivity may be due to the different mechanisms of sample analysis. Cytosponge-TFF3 employs immunohistochemistry while the Esoguard essay relies on methylation status of cyclin A1 and vimentin. Further, clinical trials utilizing Cytosponge reported outcome of long segment BE and have excluded short segment BE patients in reports of device diagnostic accuracy. Our study reported on diagnosis of short and long segment BE jointly. If all cases of short segment BE were excluded from this study population, disease prevalence would be 5.5% (95% CI 2.0, 11.5). Test sensitivity would be equal to 83.3% (95% CI 35.9, 98.6), and negative predictive value would be 98.6% (95%CI 92.1, 99.8).

The positive predictive value of this test, 32.5%, was based on the prevalence of BE among the study participants at our VA hospital which was equal to 12.9%. This number is higher than expected, yet still consistent with what was reported previously in patients with GERD screened for BE. In our prior study where transnasal and capsule endoscopy were used for BE screening, prevalence of BE was 10.6%^19,20^. Increased prevalence of BE among Veterans has been also observed by Nguyen et al^21^ who observed that it was almost 3-fold higher compared to the general US population, 8.6% vs. 3%. General population prevalence of BE has been previously reported in a systematic review and meta-analysis of 49 studies which estimated prevalence of this condition based on presence or absence of risk factors^22^. Overall pooled prevalence of BE in low-risk populations was 1.1%. The rate of BE increased by 1.2% for each additional risk factor studied. Prevalence yielded for our study population may be enriched due to our enrollment criteria which included patients with diagnosis of chronic reflux with additional risk factors for BE, and recruitment among screening accepting individuals most of whom were referred from primary care or GI clinics for colorectal cancer screening. Referral bias and participation bias may have influenced our estimate of BE prevalence among veterans.

The mean number of risk factors reported in this study was 4.1 (SD 0.8). Six of the study participants stated family history of esophageal cancer. One of the patients who was diagnosed with esophageal adenocarcinoma did report family history of upper GI tract cancer. Available data suggests that family history is the strongest predictor of BE diagnosis, as prevalence of BE among those with family history was 23%. This points to high priority of pursuing screening in patients with family history of the condition, followed by patients who share multiple risk factors. Lack of screening in patients with multiple risk factors for the condition is highlighted in our population sample and has been similarly reported by others.

We reported that Esocheck did not collect enough cellular material for methylation analysis in 10.5% of individuals which is higher than what was previously reported in studies utilizing Esoguard®^23-25^. The method of DNA extraction and amplification was updated by Lucid Diagnostics during the study period in 2022. Prior to this update, the quantum insufficient for analysis (QNS) rate was 20%. When extraction methods were optimized, the QNS rate decreased to 3/76 (4%), consistent with data from prior reports. Most QNS samples were collected early in the study period. Learning curve of test administration could have contributed to the high early QNS rate, as test administrators have to appreciate the tactile sensation when balloon opposes the GE junction wall. We performed additional analyses in which QNS results were considered positive, in the sense that an EGD would be required, and sensitivity and PPV were not diminished.

The STAI-6 questionnaire was used to assess pre- and post-procedure levels of anxiety in six areas (Figure 2). The STAI-6 score range is 20-80 with higher scores indicating more severe anxiety. The six-item questionnaire does not differentiate between mild, moderate or severe levels of anxiety. Scores for STAI-6 in this study ranged from 20 to 60, with a left skewed distribution. Results in this study of veterans showed that most of the domains that constitute the STAI-6 score were unchanged before and after the procedure. The composite STAI-6 scores were relatively unaffected by completing the Esocheck test. Participants did report statistically significant decrease in sense of worry following administration of Esocheck. The use of this validated questionnaire affords comparison of procedure related anxiety with BEST-3 trial participants^26^. When compared to screening with Cytosponge, the total STAI-6 scores were higher for Esocheck before the procedure, but scores in both studies fell to the same level of after completion of the screening test; below the anxiety threshold demonstrating comparable tolerance for the devices.

Good tolerance and acceptability of non-endoscopic screening was not surprising and supports recent findings of a discrete choice experiment in performed in Netherlands in which respondents expressed preference for noninvasive screening modalities over endoscopic and capsule based techniques when hypothetical test sensitivity and specificity were over 80%^27^. Low test sensitivity had the highest negative impact on screening participation. Esocheck sensitivity of 92.9% (95% CI 66.1, 99.8) is therefore reassuring as acceptable for screening based on previously reported patient preferences.

Calculations regarding effectiveness of 2-step screening strategy afforded by Esocheck indicate that the burden of screening would be reduced by at least 53%, likely more with a lower QNS rate than observed here. Given that the size of screen eligible population in the United States varies based on differing criteria from 4 published professional societal guidelines and ranges anywhere from 19.7 million to 120.1 million^28^, decreasing numbers of EGDs that need performed for screening by 62% could significantly decrease the burden of screening upper endoscopies. A formal cost-effectiveness analysis is being conducted and will be published separately.

A strength of this study is its prospective design, preventing conflated specificity by prohibiting participation of those who have already received an endoscopy. Another strength of this study is that those performing the endoscopy were blinded to the result of the EsoCheck test. The limitation of the study is the elevated prevalence of BE and EAC in the veteran population which may be caused by referral, selection and participation bias. Secondly, sensitivity and specificity of screening upper endoscopy was presumed to be 100%. This assumes perfect recognition of non-dysplastic and dysplastic BE by endoscopists and pathologists alike which is not reflective of clinical practice. Retrospective analysis from DeBakey VA showed that approximately 30% of veterans diagnosed with EAC had a prior upper endoscopy within a year diagnosis yet still developed interval cancers, highlighting the limitations of setting EGD as a gold standard test for BE screening^11^. Our study also highlights the importance of screening for BE and EAC in at risk veterans, as the condition was prevalent among those with risk factors.

Further studies can explore barriers to screening for BE and EAC to determine the role of EsoCheck/Esoguard in increasing access to care as the test can be performed unsedated in outpatient clinics. Because 40% of patients with esophageal cancer present without prior history of gastroesophageal reflux symptoms, the efficacy of EsoCheck/Esoguard needs to be explored in a non-GERD population of patients who have multiple risk factors for EAC. Furthermore, modeling studies can help to delineate who would benefit from screening, how often the screening needs repeated, and whether there is an age when screening should be discontinued. Furthermore, while the current cost of the Esocheck/Esoguard is high and the test is not covered by many insurance plans, continued advances in automation of the Esoguard assay should bring the cost down over time and increase affordability. Modeling studies can be used to determine the cost effectiveness of screening relying on Esocheck/Esoguard, appropriateness of screening for different patient groups, and the frequency at which they should be done by assessing quality of life years gained, as has been explored with other non-endoscopic devices^16^. Future research can survey patients and medical providers regarding their knowledge of BE screening and surveillance and query providers’ perceived capacity to implement EAC screening.

In summary, the EsoCheck device yielded a high sensitivity of 92.9% which makes it an excellent candidate for a first line screening test to rule out BE/EAC. Its specificity of 72.2% indicates a lower than desired true negative fraction, an area of possible improvement for future iterations of the Esoguard assay. Anxiety levels among participants who opted in for screening was notable but reported overall tolerance scores were excellent. Given the increasing prevalence of EAC, rising awareness of undiagnosed BE in asymptomatic individuals, and improved effectiveness of ablative and endoscopic resection techniques available to patients with early stages of disease, this screening platform opens the window to improved prognosis for EAC by increasing access to minimally invasive, well tolerated office-based testing.

## Data Availability

All data produced in the present work are contained in the manuscript.

## Notes

### Competing Interest Statement

Dr Amitabh Chak has a patent on use of methylated vimentin for detection of GI cancers and a patent on Esocheck/Esoguard device. The contents presented here do not represent the views of the U.S. Department of Veterans Affairs or the United States Government.

### Funding Statement

This study was funded by DOD Award W81XWH2110586.
Study is registered at Clinicaltrials.gov NCT05210049.

### Author Declarations

Ethics committee/IRB at the Louis Stokes VA Medical Center and USA MRDC Office of Human and Animal Research Oversight gave ethical approval for this work.

